# Pilot Study to Determine the Efficacy, Feasibility, and Impact of Storage Conditions on At-Home Blood Collection Kits for Proteomic Studies

**DOI:** 10.1101/2025.05.14.25327396

**Authors:** Caroline Scranton, Xiaoxiao Sun, Dominic Rodriguez, Kristen Pogreba-Brown, Erika Austhof, Caitlyn M McFadden, Victoria Obergh, Kerry K Cooper

**Affiliations:** School of Animal and Comparative Biomedical Sciences, The University of Arizona, Tucson, AZ, USA; Department of Epidemiology and Biostatistics, The University of Arizona, Tucson, AZ, USA; BIO5 Institute, University of Arizona, Tucson, AZ 85719, USA

**Keywords:** Proteomics, capillary, shipping, blood collection, storage

## Abstract

**Background:** At-home blood collection kits have the potential to greatly increase the efficiency of blood collection for diagnostic or research purposes by reducing the cost and burden on participants, researchers, or physicians and eliminating the need for a phlebotomist, specialized equipment, and on-site processing. These kits have shown to be effective for studying specific blood metabolites and proteins, but for analyses targeting the entire proteome, their effectiveness is unknown.

**Methods:** For this pilot study, data on human serum proteome was compared when blood was capillary-collected with a Tasso+ device (Tasso Inc.) versus the gold-standard venous samples drawn by a trained phlebotomist. Analyses were conducted using the SomaScan 7K assay (SomaLogic Inc.), which assesses the levels of nearly 7,600 serum proteins. Additionally, duplicate Tasso+ blood samples were also subjected to a variety of pre-processing storage temperatures and times to mimic the effects of shipping samples from participants on the serum proteome compared to baseline samples.

**Results:** Minimal differences were seen between the serum proteome results of capillary and venous blood for all participants. Delays in processing of greater than 48 hours led to large changes in detected protein levels throughout the serum proteome, while lower holding temperatures (refrigeration at ≥4°C) pre-processing decreased the amount of change in the serum proteome.

**Conclusions:** Overall, it was determined that when processed immediately, capillary blood gives similar results to venous blood, while minimizing the time (≤48 hrs) and temperature (≤4°C) can minimize the serum proteome changes in samples collected by at-home blood collection kits and detected by the 7K assay.

## Introduction

At-home blood collection methods have become increasingly popular for medical testing, surveillance, and research, particularly in light of the SARS-CoV-2 pandemic, which added increased barriers to healthcare and participant-based research (1). However, because at-home blood collection kits must be mailed in for analysis, they must be able to stabilize the blood in the presence of varying temperatures over extended periods of time. Extreme temperatures and shipping delays may significantly affect blood samples and potential analysis results. Therefore, it is critical to understand how different conditions impact samples and analysis results for those collected outside of the clinic setting to effectively assess the results compared to traditional collection methods.

The Tasso+ kit (Tasso Inc, Seattle, WA), an at-home blood collection kit developed in 2022, collects capillary blood as opposed to venous blood that is collected through standard phlebotomy procedures (2). Tasso+ devices have previously been used to collect blood for Anti-SARS-CoV-2 IgG antibody testing for several studies, which found that the levels of IgG in samples collected using venous blood versus capillary blood (collected with Tasso+ kits) were quantitatively similar (3–5). Additionally, Tasso+ kits have been shown to keep blood stable after extended time and temperature conditions (to simulate shipping) in several studies, when looking at specific IgG antibodies, ions, proteins, and vitamins (4, 6). The effects of pre-analytical variation, including collection tube type and use of stabilization agents on the proteome are variable, but also has not been widely explored with high-throughput proteomics (7, 8). With the development of high-throughput proteomic assays, an expansive amount of information about an individual’s proteome can be detected from a small volume of serum, including disease biomarkers, metabolic issues, and more (9). Proteins vary in stability outside of the body (10), and while ideally every blood sample would be collected and processed immediately, this is often not feasible in large, population-based studies and poses a significant barrier to data collection. However, technologies such as the Tasso+ kits may be able to overcome these study limitations (11, 12). Evaluation of the effectiveness of these kits potentially allows for an expansion of at-home sample collection, allowing researchers and physicians to overcome significant barriers imposed by traditional blood collection methods (1).

The SomaScan 7K assay (Somalogic, Boulder, CO) is a proteomic assay that uses single-stranded DNA aptamers (SOMAmers) with modified uridine residues specifically designed to bind to nearly 7,600 different proteins to measure protein levels in serum (13). The assay uses DNA hybridizing microarrays with the SOMAmers forming complementary structures to their corresponding protein (14). The relative levels and the dissociation rate of each SOMAmer detected in the sample is directly related to the level of a specific protein in the serum (14, 15). Next-generation proteomics assays like the SomaScan 7k allow for a deeper look into the physiology and metabolic patterns of individuals – this is a more efficient, albeit high-cost, method to look at a wide range of proteins, compared to traditional methods like mass spectroscopy or immunoassays. The ability to identify key biomarkers of acute and chronic diseases and a method of linking broad disease symptoms to specific biological imbalances make next-generation proteomics a valuable tool in health-based research and diagnostics (9).

The aim of this pilot study overall is to determine the efficacy of using Tasso+ kits in high-throughput proteomics, specifically (1) assess whether the kits can keep blood samples stable over extended periods of time (i.e. shipping from participant’s home to laboratories); (2) assess impact of storage temperature on the blood samples (i.e. various shipping temperatures) as a stable low temperature cannot be guaranteed when shipping samples, even with ice packs or similar measures; and (3) assess whether the capillary blood collected by the kits has a proteome equivalent to venous blood collection. Additionally, the study also assessed the ability to use at home blood collected with the Tasso+ kits for next-generation proteomics, which greatly extends the reach of these new and continuously improving techniques. If effective, these kits have the potential to reduce the burden on researchers, physicians, and participants by collecting samples in a less-invasive manner, particularly where participants are impeded by barriers such as distance from a clinic, ability to travel either due to resources or health status, time during traditional work hours needed for a phlebotomy appointment, distrust of traditional healthcare settings or even fear of needles (16). This pilot study will provide context for the feasibility of using Tasso+ kits for proteomics in a large cohort study. This will also expand the reach of high-throughput proteomics, allowing researchers and doctors to surpass significant barriers on blood sample collection and gather more data on an improved scale.

## Materials and Methods

### Participants and Setting

This pilot study involved four participants, each providing 8 blood samples each. Due to funding restraints, more participants and blood samples were unable to be collected. Participants varied in age and sex, with one male and one female participant in their 40s and one male and one female participants in their 20s. This study was approved by the University of Arizona Institutional Review Board (Protocol: STUDY00003431) and all participants provided informed consent prior to participation. This study was completed as part of a pilot to test biospecimen collection and testing procedures for a larger cohort study, the Arizona CoVHORT-GI (17). A main motivation factor in conducting this study was to assess the stability of biospecimen collection during the extreme temperatures common to Arizona in the summer months.

### Venous blood collection

Figure 1 outlines a workflow of sample collection and processing. The four participants each provided one venous sample of roughly 5 ml (collected in a 10 ml BD Vacutainer serum-separating tube (SST)). Venous samples were collected by The University of Arizona’s Clinical and Translational Science Research Center phlebotomy team at Banner University Medical Center (Tucson, AZ) immediately prior to Tasso+ sample collection. Samples were immediately put on ice and brought to the laboratory and processed as described below.

**Figure 1.**
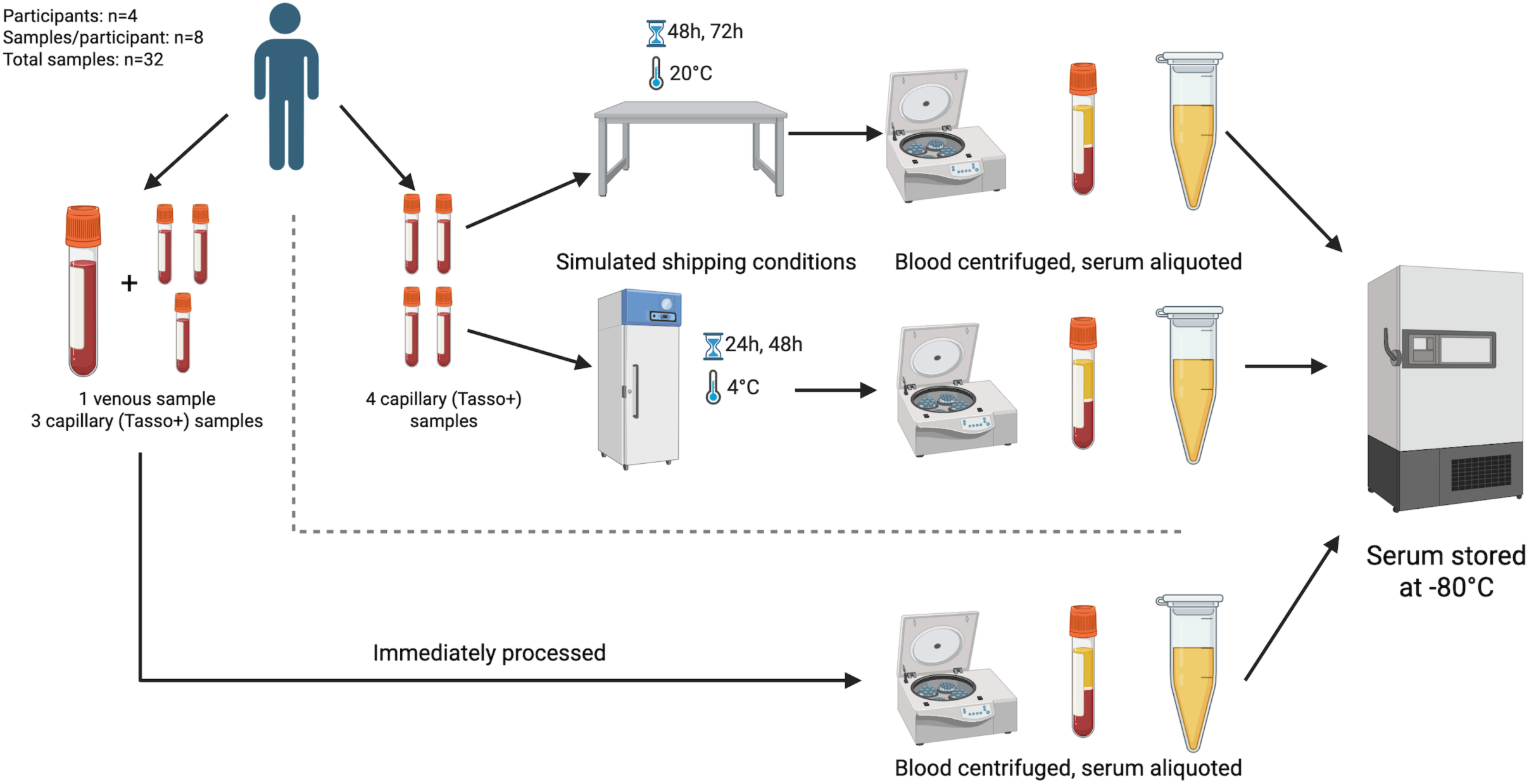
PCA plot of all samples. The principal components analysis done on the whole proteome of all 32 samples generated during the study was plotted. Samples significantly clustered by participant (p<0.001), time (p<0.001), and temperature (p<0.04) using a PERMANOVA test, but not by collection method (p=0.136). As samples were subjected to pre-processing conditions the proteome began to differ, as shown by the increased distances on the plot between the samples, however those subjected to the same conditions (i.e. storage at 4°C) were more closely related to each other than the rest of the samples, suggesting the impact of time and temperature impacts the proteome consistently among the participants.

### Tasso blood collection

The Tasso+ Kit is a self-contained at-home blood collection kit containing all of the necessary materials to collect roughly 500 μl of blood from a patient’s upper arm, thigh, or heel (18). The kit, which is classified as a class II lancet is compatible with a variety of collection tubes allowing for different reagents to be used, including blood-stabilizing reagents that potentially allow for transport and storage at room temperature (18). Tasso+ kits were chosen for this study due to previously documented collection success and use in other studies looking at components of blood (3, 4, 18). Eight capillary samples from each participant were collected with Tasso+ kits with SST tubes, according to the manufacturer’s protocol. Some samples were collected simultaneously from each participant due to the high number of collections and to minimize time between collections. The majority of samples were taken from the upper arms of the participants (both right and left side); however, several were taken from the thigh when space on the upper arm ran out. Both Tasso+ and venous blood samples were collected on the same day over the span of less than 3 hours.

### Sample Treatment

Samples were subjected to different pre-processing time and/or temperature treatments, as outlined in **Table 1**. These parameters were chosen to represent realistic shipping conditions and timeframes, as participants can collect their samples and store them either at room temperature (20°C) or in the refrigerator (4°C). Additionally, refrigeration of whole-blood samples has variable effects on protein concentration depending on the protein(s) of interest, so this was tested to see the impacts on a larger amount of the proteome as a whole (19). Overnight shipping of the samples would lead to a shipping time of approximately 24 hours, however delays in shipping by participants or delivery may result in longer periods of time prior to processing. Additional samples were collected using Tasso kits and stored at 55°C (representing a delivery vehicle in the summer of Arizona, United States) and at -20°C (frozen), but were excluded from the study as the blood had extensively degraded and such low quality that it would not pass the SomaScan 7K assay quality check. Additionally, there were cost limitation on testing a wider array of samples, so the time/temperature conditions were chosen to represent a range of realistic shipping conditions.

**Table 1.** Blood samples collected from each participant and tested with SomaScan 7K assay.

| Blood Draw Type | Temperature (°C) | Time (hours) |
| --- | --- | --- |
| Tasso | 20°C (room temperature) | 0 |
| Tasso | 20°C (room temperature) | 0 |
| Tasso | 20°C (room temperature) | 0 |
| Tasso | 20°C (room temperature) | 48 |
| Tasso | 20°C (room temperature) | 72 |
| Tasso | 4°C (refrigerated) | 24 |
| Tasso | 4°C (refrigerated) | 48 |
| Venous | 20°C (room temperature) | 0 |

### Sample processing

After subjecting samples to different treatments or controls (processed immediately), the serum-separating tubes were centrifuged 1,400 x g for 2 minutes. When removing sera from the SST tubes, roughly 5 μl of serum was left in the tube to ensure that no SST gel was removed as it would contaminate the serum. Serum from each sample (130 μl) were aliquoted into a 2 mL safe-lock Eppendorf tube (Eppendorf, Germany) as per request by SomaLogic and were stored at - 80°C until shipment from Tucson, Arizona to SomaLogic Inc in Boulder, Colorado. The remaining serum was stored in a 1.5 mL Eppendorf tube at -80°C for later use in the Cooper Laboratory.

### SomaScan 7K assay

The SomaScan 7K assay was chosen to assess many aspects of the proteome at once to look at the overall impacts of time and temperature, rather than looking at specific proteins, particularly in the context of using blood for biomarker discovery where specific proteins of interest are unknown before analysis. All samples were shipped to SomaLogic in Boulder, CO, and upon receipt were subjected to quality control assessment. All samples for this study were determined to be of sufficient quality for the assay. The SomaScan 7K assay was performed by SomaLogic as described in their technical note, which uses nucleotide aptamers, created to match nearly 7,600 unique proteins with high specificity, to indirectly measure the concentration of different proteins in the serum using a DNA microarray (15). Any remaining serum from each sample sent to Somalogic that was not used in the SomaScan 7K assay was destroyed. Data generated from the SomaScan 7K assay included the normalized measurements of 7,596 proteins for each sample, SomaLogic’s calibrators, quality controls, and buffers are all reported in relative fluorescent units (RFU).

## Data analysis

Data returned from SomaLogic was imported into R studio version 4.3.1 using packages readxl and openxlsx (20–22). The data was analyzed using the R package limma (Linear Models for Microarray and RNA-seq Data)(23). Raw RFU data first underwent a log2 transformation using the R package dplyr, as is standard with microarray data (23, 24). Next, linear models were fitted to the data, and proteins that were significantly impacted in each model (determined via empirical Bayes statistics) were identified. Significantly impacted proteins were determined as those proteins with an adjusted p-value less than or equal to 0.05 after a false discovery rate adjustment, which were sorted into KEGG (Kyoto Encyclopedia of Genes and Genomes) pathways using the enrichKEGG function in the R package clusterprofiler (25). SomaLogic raw data, processed data, and all code used for data analysis models, sorting of significant proteins into pathways, and data visualization can be found on GitHub: https://github.com/carolinescranton01/AtHomeBloodCollection-Proteomics_Analysis

1. **Sample-by-Sample Comparison.** Each participant had three baseline Tasso+ samples (processed immediately after collection) collected, which were compared to all the other participant samples using univariate linear models. The models (further referenced to as ‘model type A’) had the structure: RFU ∼ β + Group, where protein levels were measured in relative fluorescent units (RFU), beta is the intercept, and ‘Group’ was used to distinguish between the three baseline Tasso+ samples and the fourth non-baseline sample. This model was used to determine the difference for each participant individually between capillary and venous blood, storage times, and storage temperature compared to baseline.
2. **Venous versus Capillary Analysis**. A multivariate model was created to compare the capillary and venous samples across all four participants at once (‘model B’). Model B contained two parameters in addition to the intercept, blood type (‘Type’, meaning capillary (from Tasso+ kits) or venous (traditional blood draw), which was dummy variable coded as 0 = capillary and 1 = venous, with capillary considered as the reference) and participant (‘Participant’, functioning as a dummy variable to group participants together). The four samples from participant 1 had a value of 1, participant 2 had values of 2, participant 3 had values of 3, and participant 4 had values of 4, and the model was structured as RFU ∼ β + Type + Participant. The ‘Participant’ variable was fixed as the actual variation in protein levels between individuals and within the set of an individual’s samples was not analyzed, but the inherent variation needed to be accounted for in this model (as well as all following models with a ‘Participant’ variable).
3. **Impact of Storage Temperature and Time.** Model C contained three parameters in addition to the intercept and was structured as RFU ∼ β + Time + Temperature + Participant. This model was fitted to only the Tasso+ collected blood sample data. The parameters ‘Time’ and ‘Temperature’ correspond to the storage time (in hours) and temperature (in Celsius) of the individual samples. The ‘Participant’ parameter was identical to that in model B. This model incorporates the two target variables (time and temperature), as well as participant to account for inherent variation between individuals, and was used to determine the effects of each variable in respect to changes in temperature. To assess the impact of time-to-processing, model C was used to determine the effects of time-to-processing on the detected proteome when accounting for changes in temperature and variation between individuals.

## Data Visualization

Significantly different protein detection levels based on each model’s parameters were plotted using the R packages ggplot2 (26) and patchwork (27) as bar graphs by the log2-fold change in detection. Proteins were sorted by KEGG subcategories (of which there are 48 within the six KEGG categories looked at in this study) and color-coded by overarching KEGG categories (metabolism, human diseases, organismal systems, cellular processing, environmental information processing, and genetic information processing)(28). Points representing individual proteins (which were often sorted into two or more KEGG subcategories, resulting in several points with different X values but the same Y value) were plotted on the bar graph, as well as box-and-whisker plots (26). Box-and-whisker plots show the inter-quartile range (25^th^ to 75^th^ percentiles) of log2-fold change for that KEGG subcategory with a black line for the median, and whiskers extend 1.5 times the interquartile range in both the positive and negative direction (26). Additionally, Venn diagrams were created to show which proteins overlapped between participants when looking at changes in detection, using model A. Venn diagrams were created with an online tool by the VIB-UGent Bioinformatics and Evolutionary Genomics department (https://bioinformatics.psb.ugent.be/webtools/Venn/). Principal components analysis was done on the whole proteome of the samples, generated using the covariance matrix of the data (using the prcomp function in base R, and PERMANOVA tests were done using the adonis function the R package vegan (29)), showing the variation between samples overall. Plots were made using the R packages ggplot2, ggfortify, and ggrepel (30, 31).

## Results

### Serum quality for SomaScan 7K assay

SomaLogic provides a quality statement with all SomaScan 7K assay results to inform researchers the quality of their samples, both during the run and after normalization. All 32 samples in this study passed every quality check, signifying that despite exposure to adverse temperatures over extended periods of time, the serum retained enough quality to be assessed by this assay. However, 179 of the 7,596 assessed proteins in the SomaScan 7K assay were flagged as the ratios of the measured protein in the quality control sample compared to the study sample were outside of normal range for all samples (**Supplementary Figure S1**).

### Initial analysis

The entire proteome from each sample was analyzed with a principal components analysis using the covariance matrix of the data (7,596 proteins for all 32 samples; n=243,072 data points) to assess the overall variation in each sample. Several clusters could be identified based on both time and temperature (**Figure 2A**). Over the entire dataset, there was significant clustering by time-to-processing (PERMANOVA, p<0.001; **Figure 2A**), temperature (PERMANOVA, p<0.04; **Figure 2A**) and participant (PERMANOVA, p<0.001; **Figure 2B**), but not by collection method. Univariate models comparing every non-baseline sample per participant to the baseline samples per participant (models included only one variable to separate the baselines from the other sample, identically to model A) was conducted to assess the conditions that resulted in the lowest number of impacted proteins compared to baseline samples. The lowest number of impacted proteins on average was identified for samples that were stored at 4°C for 24 hours, which, across the four participants, ranged from 584 to 2,831 proteins (7.69% to 37.27% of the measured proteins) impacted by the conditions or an average 1,655.5 or 21.79% (**Supplementary Table S1**). The number of overlapping proteins with significantly altered detection levels per the five non-baseline sample types per participant are presented in Venn diagrams, with 52, 271, 0, and 287 for participants A through D respectively (**Supplementary Figure S2A-D**). Only one protein, neurophysin 1 (oxytocin carrier protein) had significantly different detection levels in all samples from participants A, B, and D (Participant C had no significantly altered proteins in all samples; **Supplementary Figure S3**). Overall, 202 identified proteins were not altered in any of the samples. The protein significantly altered in all samples is listed with its function in **Supplementary Table S2**, and those not significantly altered and their functions (some of these proteins have multiple functions and are listed several times) are listed in **Supplementary Table S3.**

**Figure 2.**
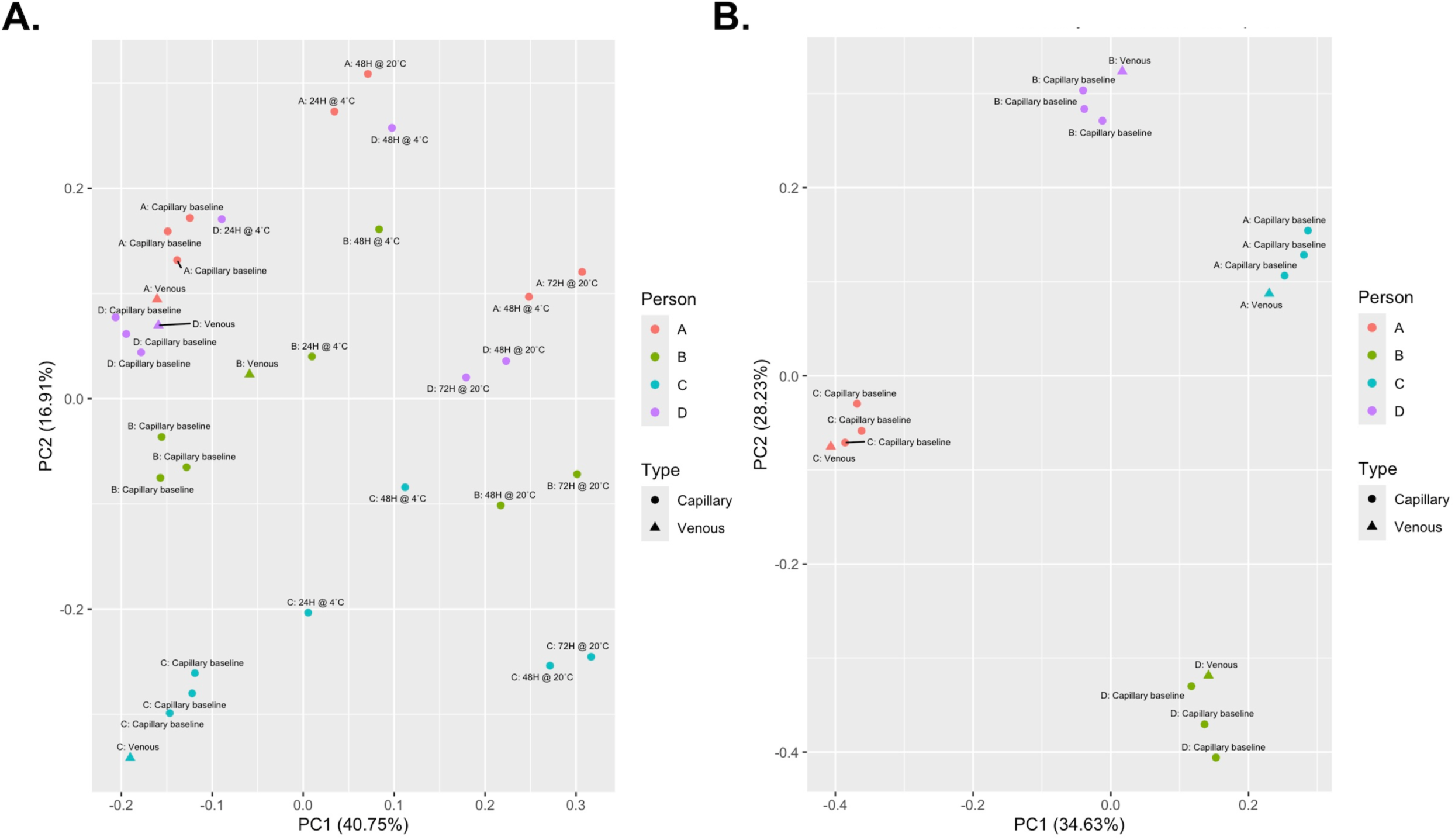
PCA plot for proteome of capillary versus venous blood samples. Each participant’s three capillary baseline samples (which were collected and immediately processed) and venous sample (also immediately processed) were analyzed using a principal coordinates analysis. Samples are significantly clustered by participant (PERMANOVA, p<0.001) and not by collection type (PERMANOVA, p=0.328), suggesting that the variation in the proteome between samples is due to individual variation rather than collection type.

### Capillary versus venous blood

Comparison of the proteome of the venous blood sample to the capillary samples for each of the four participants found that venous and capillary protein were similar to each other for each participant when processed with the same temperature and time conditions. The venous and capillary samples were significantly clustered by participant using a principal components analysis (PERMANOVA, p<0.001), but did not cluster by collection method (PERMANOVA, p=0.328; **Figure 2B**), therefore collection method was not significantly impacting the overall proteome. While not statistically significant, there were still differences in protein detection between the collection methods, as participants A through D had 89, 77, 1, and 300 proteins with decreased detection and 19, 393, 0, and 330 with increased detection respectively. The extent of that impact (in log2-fold change) varied between participants and was significantly different between almost all participant comparisons (A vs B, A vs D, B vs C, B vs D, and C vs D were significant, but A vs C was not) (Kruskal-Wallis with Dunn correction, p=0.183; **Supplementary Table S4**). The proteins perform diverse functions in the body, but the KEGG categories with the most noticeable differences were human diseases, organismal systems, and metabolism. (**Supplementary Figure S4).** Of the proteins with significantly different detection in the venous compared to the capillary blood, none were shared among all four participants (**Figure 3**). For individuals B and D, the majority of proteins with significantly impacted detection levels in the venous sample were unique to those participants (62.01% and 67.23% respectively), whereas 57.01% of participant A’s impacted proteins and 100% of participant C’s were shared with participant D. Only 14 proteins were shared between participants A, B, and D (none of participant C’s proteins were shared with all A, B, and D), suggesting that individual variation may play a greater role in determining what proteins are detected at significantly different levels from the two blood collection methods. Uniprot IDs and functions for the proteins with significantly impacted detection in Model B are listed in **Supplementary Table S5**.

**Figure 3.**
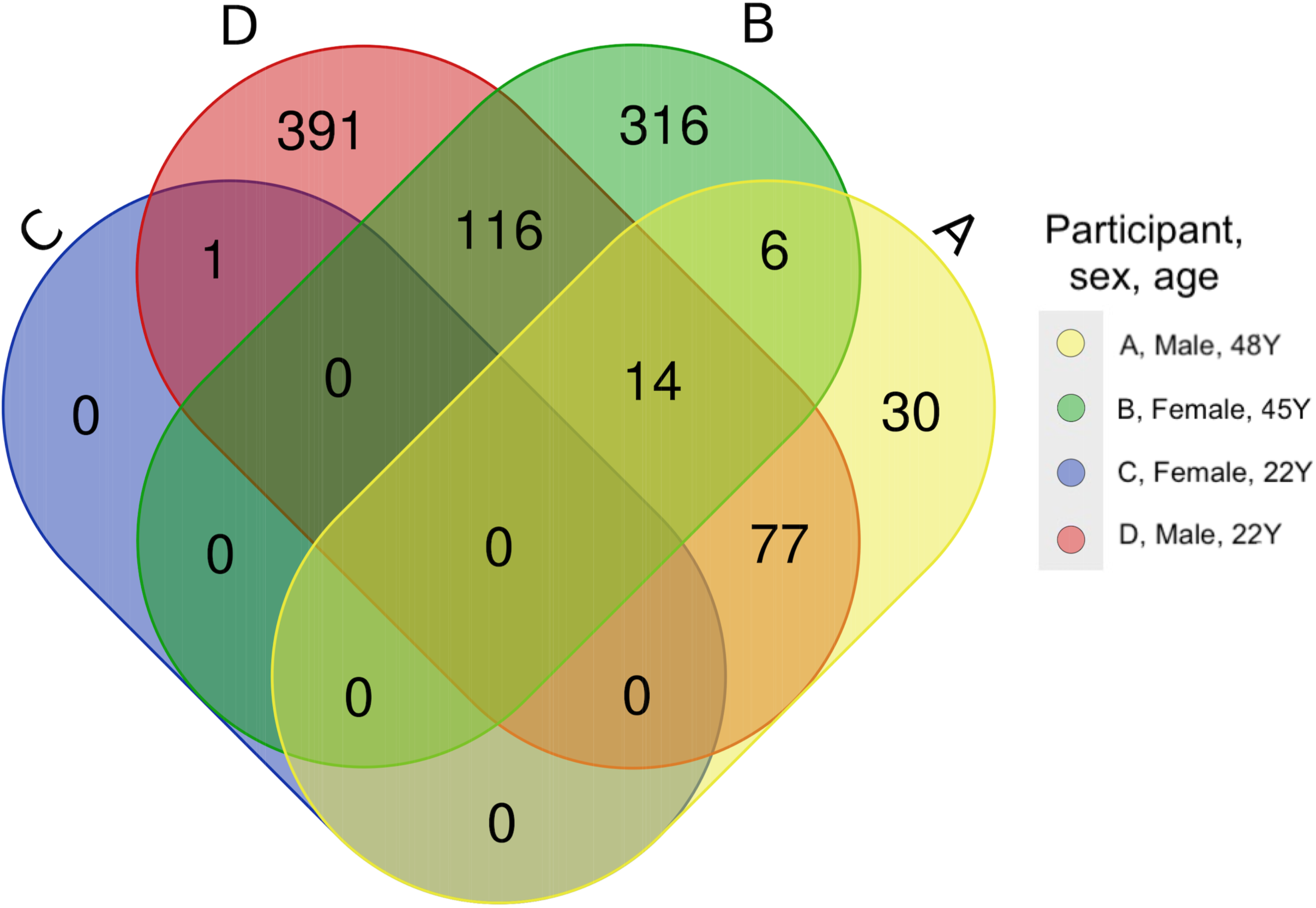
Capillary versus venous Venn diagram. The proteins which were significantly altered in detection for each of the participants individually were compared, and no proteins were altered in all four models. Panel letters A through D correspond to participants A through D. The overlap in impacted proteins varies between participants, with the majority of impacted proteins being unique to participants B and D, shared between B and D, or shared between D and A.

The multivariate model (Model B), which looks at the impact of collection method using all four participant’s data, found that 165 proteins in 37 of the 48 KEGG subcategories (not including N/A) were detected at significantly different levels when blood was collected from the veins compared to the capillaries (2.17% of the measured proteins). Many of the proteins had slightly lower detection in the venous samples compared to the capillary samples. The log2-fold change in protein detection levels ranged from -3.77 to 0.82 units, however the majority of change ranged from -2.5 and 0.1, with an overall average of -0.612 units (**Figure 4**). The protein with the largest decrease was P31947 (14-3-3 protein sigma, a signaling protein (32)), which decreased in detection by 3.77 log2-fold, while P16118 (6-phosphofructo-2-kinase, involved in glycolysis (33)) increased in detection by 0.728 log2-fold. When compared to the variation by the individual participants, the extent to which proteins increased in detection is smaller and more consistent across KEGG categories for the participants together, while the proteins that decreased in detection are similar to those shown for participants B and D (**Supplementary Figure S4)**. This suggests that while the detection of some proteins by this assay may be altered by blood collection method, when looking at the study participants together and controlling for individual variation, this only affected 165 of the 7,596 measured proteins. However, this impact was inconsistent across individual proteins and was distributed across proteins with a variety of functions.

**Figure 4.**
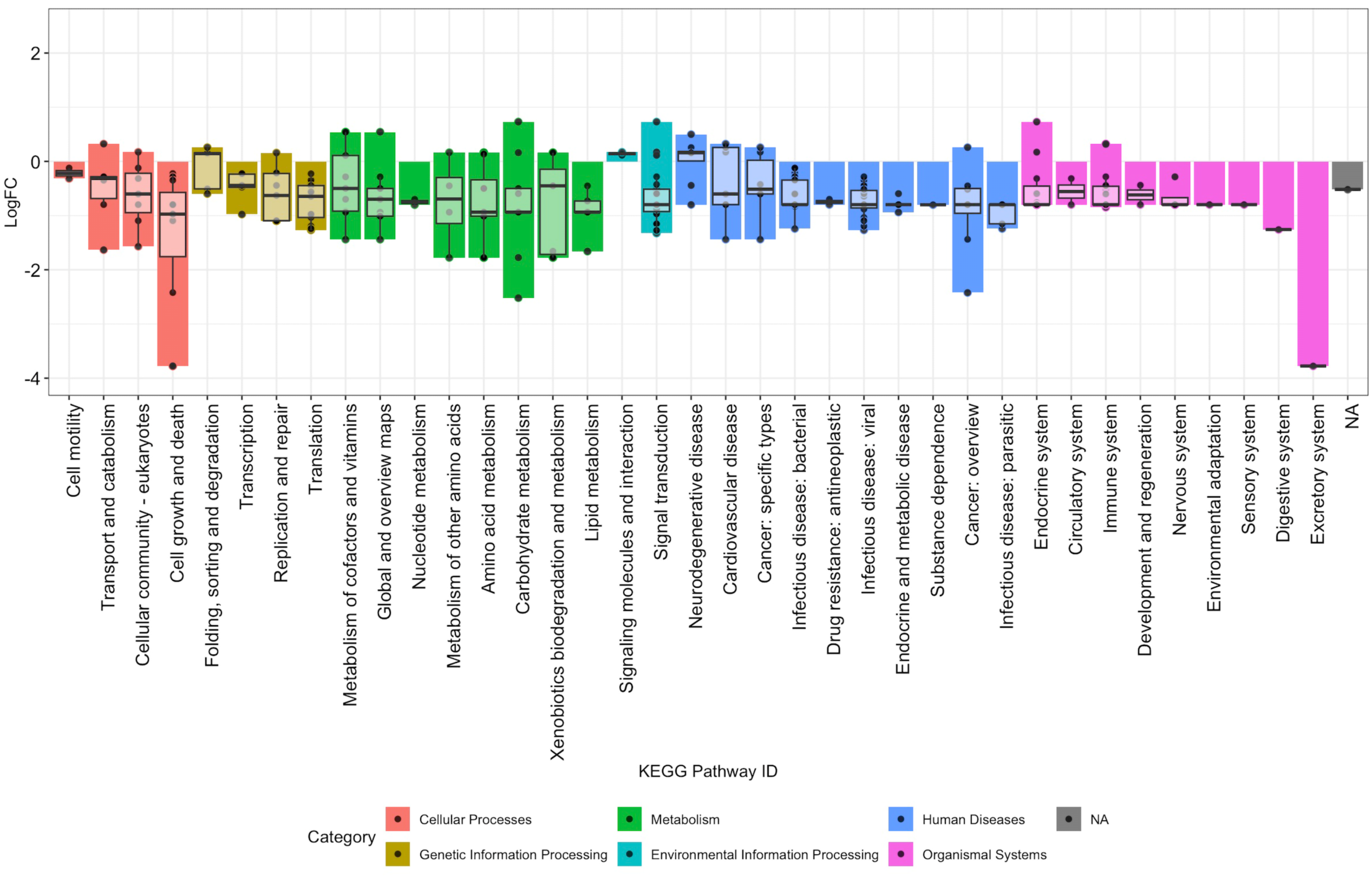
Pathways impacted by collection method, from Model. **B.** The log2-fold change in protein detection for significantly impacted proteins under Model B (multivariate model which looks at the difference between capillary and venous blood, using data from all participants and considering variation between the individuals) was plotted according to KEGG subcategory (x axis) and colored by KEGG category. A total of 165 proteins were impacted and are shown on the plot, with log2-fold change ranging from -3.77 to 0.82.

### Impact of pre-processing storage temperature

To assess the impact of temperature on the measured proteome of the blood, Model C was utilized, which compared the samples subjected to adverse pre-processing conditions against the capillary baseline samples for all four participants at once. Model C detected 3,376 proteins sorted into 46 of the 48 KEGG subcategories that were significantly impacted by temperature, with time and individual/participant variation being considered. The change in the protein detection levels was variable across proteins and KEGG subcategories, ranging from -0.13 to 0.23 units of log2-fold change. The subcategories with the highest rates of increased detection were human diseases and environmental information processing. The variation in the interquartile range for each subcategory was quite consistent across 46 of the 48 KEGG subcategories, and was skewed slightly negative, with the largest differences between subcategories resulting from extreme values designated as outliers in the box and whisker plots (**Figure 5; Supplementary Table S6**).

**Figure 5.**
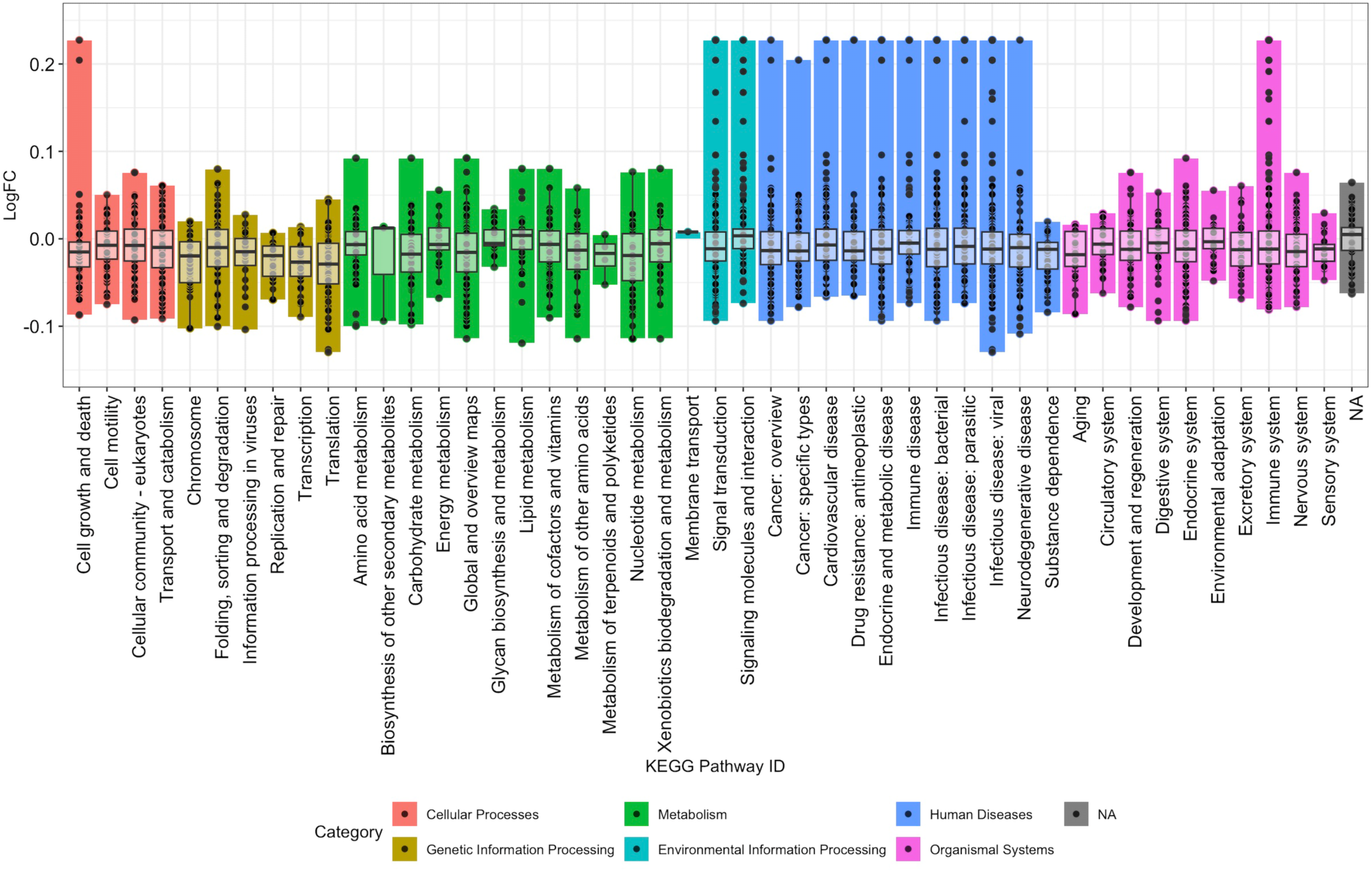
Pathways impacted by storage temperature. The proteins with significantly different log2-fold change under model C, which looks at the impact of temperature on protein detection while considering individual variation and variation in processing time when compared to the capillary baselines. A total of 3,376 proteins were significantly impacted in this model. The spread of the impacted proteins is somewhat consistent across the different KEGG subcategories, ranging from -0.13 to 0.23 units of log2-fold change, while the boxes in the box-and-whisker plots range from roughly -0.05 to 0.02 units. Certain proteins were detected at higher levels (i.e. Interleukin 6 (IL-6), UniprotID: P05231, sorted into 13 subcategories, was detected at 0.23 units of log2-fold change), but much of the data centered around no log2-fold change.

### Impact of time-to-processing

The impact of time on the detected proteome was also assessed. When looking at individual participants, after 48 hours the average number of proteins with impacted detection was 4350.5 (57.27% of the SomaScan 7K panel), rising to 5197.75 (68.42%) after 72 hours (**Supplementary Table S1)**. Using Model C, which found 5,159 proteins with significantly different detection over extended periods of pre-processing storage time, again falling into 46 of the 48 KEGG subcategories. Although a higher number of proteins were impacted by extended processing time (67.9% of detected proteins) compared to collection method (2.17%) or storage temperatures (44.4%), the log2-fold changes in detection of these significantly different proteins was lower, ranging from -0.036 to 0.105 units, with the middle 50% of the data centered around the 0.0 log2-fold change. Similar to the impact of storage temperature, storage time had the same pathways (human diseases and environmental information processing) as well as organismal systems with outliers that showed much larger increases in detection than the majority of the data (**Figure 6; Supplementary Table S7**).

**Figure 6.**
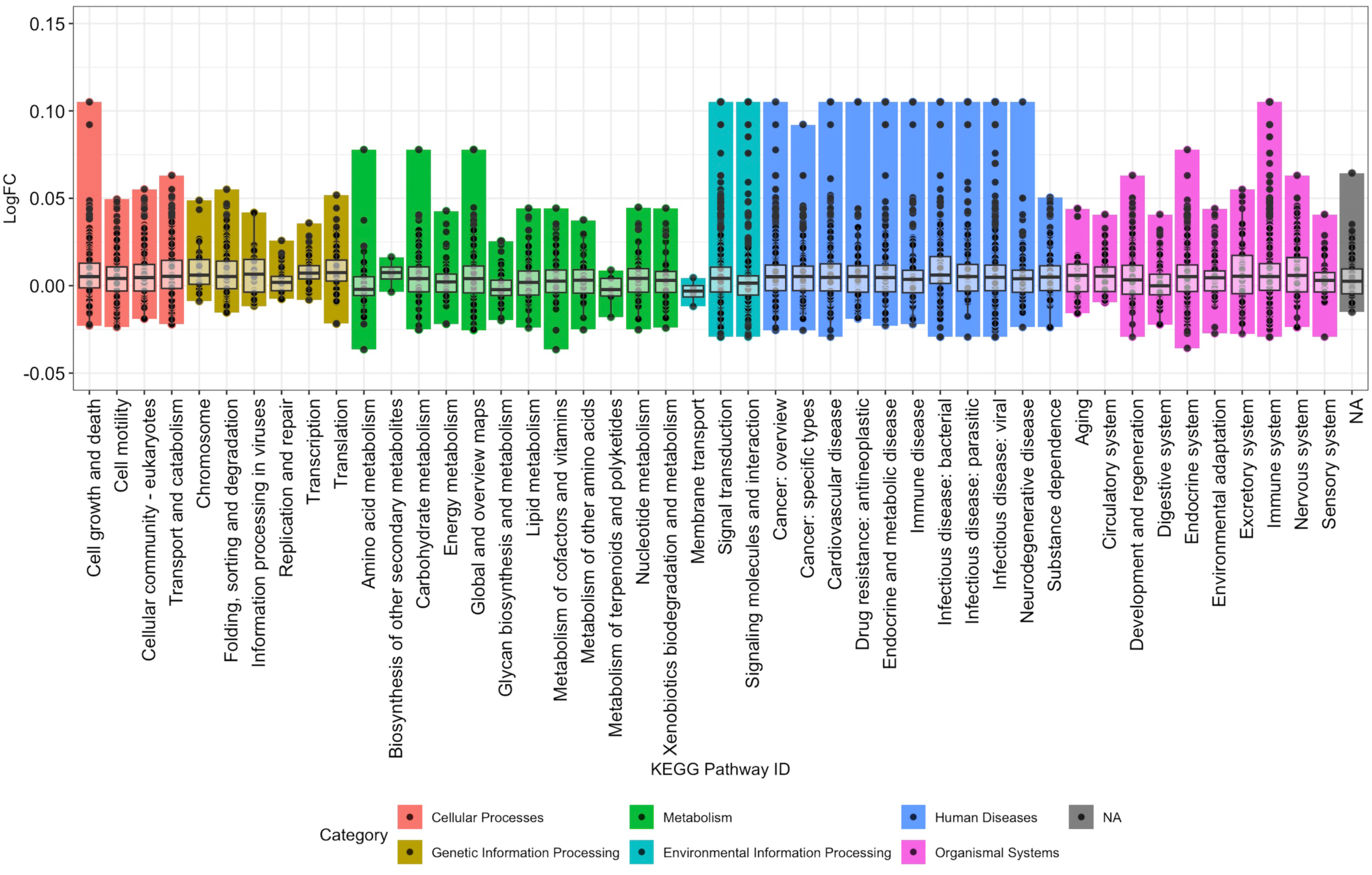
Pathways impacted by time-to-processing delays. The proteins with detection impacted significantly by time-to-processing identified through Model C to determine which proteins had been detected at different levels in the samples with extended time-to-processing compared to the capillary baselines for all four participants, while taking temperature variation and individual variation into account. A total of 5,159 proteins were impacted in this model and are represented on the figure. Similar to the impact of temperature, some subcategories have much larger log2-fold change values (the overall range is -0.036 to 0.105 units) while the middle 50% of the data is small and centered around no log2-fold change (ranging from roughly -0.01 to 0.01 units). Although more proteins were impacted by time, the extent of the change was less than that of temperature.

## Discussion

This pilot study compared the proteome of capillary blood collected with Tasso+ kits to venous blood using serum samples from four participants, which were analyzed using high-throughput proteomics. Despite some differences in protein detection levels in the venous samples and samples subjected to adverse pre-processing time and temperature conditions, all samples were suitable for analysis as determined by the SomaScan 7K assay. Other studies have found Tasso+ collected blood to be suitable (and comparable to venous blood) for analysis on a range of compounds in the blood, including for use in the OLINK high-throughput proteomics assay (measures 384 biomarkers related to inflammation)(3, 4, 7). At-home blood collection kits are convenient and reduce burden on physicians/laboratories and patients/participants, but detection of individual proteins may be impacted by uncontrolled pre-analytical variables (4, 34).

Comparing venous to capillary blood, proteomes significantly clustered by individual but not by collection method, meaning proteomic differences are likely more influenced by variation between individuals than blood collection method. Model B, which considers all participants simultaneously, found slightly lower levels of significantly impacted proteins in venous than capillary blood. Previous research has found the levels of many specific analytes in blood, such as SARS-CoV-2 IgG antibodies, hemoglobin, and clotting factors, were not detected in significantly different amounts on capillary versus venous blood, which is supported by this study (4, 5, 35, 36). Although the SomaScan 7K assay was developed for venous blood, capillary blood has provided similar results to venous blood in this pilot study, suggesting it is feasible to use capillary blood in this assay. The variation in protein detection in this pilot study is most likely explained by individual differences (which can be accounted for in analysis) rather than blood collection method, meaning that potentially random individual differences may normalize with a larger study population. Therefore, while the results of this pilot study find capillary blood is a viable option to expand the reach of large-scale high-throughput proteomic studies without impacting the quality of the data, given the small number of studies that have addressed these differences, it would not be advisable for a single study to use more than one blood collection method.

Model C assessed the impact of temperature on the proteome and found fewer significantly impacted proteins (44.4%) than when looking at the impact of time, but the range of log2-fold change of impacted proteins was larger. Storage at lower temperatures (4°C versus 20°C) led to slightly lower protein detection although some proteins, such as interleukins 6 and 8, had large increases in detection. Chang et al. found that higher pre-processing storage temperatures lead to significant changes in protein detection, while storage at ∼25°C maintained protein stability for up to 8 hours – a significant amount of time, but too short to realistically represent shipping of samples (11). Studies have found both increases and decreases in protein detection when looking at specific proteins in blood stored above room temperature (30°-37°C) which was also found in this piloty study (11, 37). Refrigerating samples prior to processing has been shown to reduce protein degradation and hemolysis in general (38). The impact of rising temperature (increasing from 4°C) over time was not tested, but one study found that a temperature rise from 4°C to 13°C over 24 hours had negligible effects on certain biomarkers in whole blood (including IL-6, which was significantly impacted in almost all models in this pilot study) compared to storage at room temperature (4). Although the effect of temperature can be mitigated in some ways, such as shipping samples on an ice pack, a stable, low temperature is not guaranteed, especially in warmer weather or if shipping delays occur. Understanding the effects of temperature on the proteome is crucial, and this pilot study and others have shown that while Tasso does not specify shipping temperatures (18), storing and shipping samples at 4°C may preserve the overall proteome to account for time it takes to ship the sample back to the laboratory, especially if this time exceeds 24 hours.

Extended time-to-processing impacted protein levels significantly when samples were stored at room temperature. Delays in the first 48 hours are contributing more greatly to changes in protein detection, as the difference between 48 and 72 hours is smaller than that of zero to 48 hours. The magnitude of log2-fold change due to extended time-to-processing was smaller than that of temperature in Model C. The study found that the same two inflammatory proteins had the highest changes for time and temperature models, P05231 (interleukin 8) and P10145 (interleukin 6). Both proteins are important diagnostic biomarkers and are naturally variable in healthy individuals, which may have also been locally elevated in the participants due to repeated blood sample collection (39, 40). Hassis et al., found increased detection of specific proteins after 96 hours of pre-processing storage at room temperature (37), while El-Sabawi et al. found changes in detection of 88.7% proteins using the OLINK high-throughput proteomics assay after storage at 4°C for 24 and 48 hours, aligning with the results of this study, particularly when looking at inflammatory proteins (7). Time-to-processing delays appear to impact many proteins in the proteome, although log2-fold detection changes are smaller than the impact changes caused by temperature or collection methods. The impact of time-to-processing should be noted if planning to use at-home blood collection methods for protein studies, as time-to-processing can be difficult to control when shipping samples, even locally.

Refrigerating blood may reduce the impact of extended time-to-processing on the proteome. The number of proteins impacted for each of the participants after 48 hours at 4°C was much lower than at 20°C. Despite efforts to control for the effects of time, storage temperature, and individual variation, it is clear that the proteome does change extensively when the blood is not processed immediately. Storage at room temperature for short periods of time (<8-12 hours) is ideal (11) and recommended by Tasso (18). While refrigeration may impact protein detection levels, this pilot study found that it is recommended if the sample must be stored for longer periods of time. For clinical studies, shipping capillary blood is feasible if the shipments can ideally be temperature-controlled (i.e. samples are shipped with an ice pack) and returned to the lab in under 24 hours to minimize variables not associated with the participant. This is particularly important in areas that experience extreme temperatures during the summer months, as samples in this study that reached extreme temperatures were degraded to the point of not being usable.

Other factors may influence protein detection in blood samples aside from blood collection method and storage conditions. Nedelkov et al. found that 18 of 25 clinically-relevant proteins had slight structural modifications, as either point mutations or post-translational modifications between the 96 participants in the study (41). Thus demonstrating that serum protein levels are inherently variable and certain proteins can vary structurally between individuals, along with protein degradation over time (particularly in extreme temperatures), both can inhibit aptamers from binding specifically to proteins thus changing the detection levels of some proteins (41) depending on the proteomic assay utilized for the study. The proteome is naturally variable throughout the day – all samples in this pilot study were collected over a short window of time (3 hours), aiding in reducing the effects of this natural variation(42). Certain immune-related proteins, such as interleukin 8 (Uniprot ID: P10145), had increased detection, potentially due to activation of the innate immune response as participants collected multiple blood samples over a short period of time from the same area of the body (upper arm, both right and left)(43) (44). Hemolysis was evident in all non-baseline samples to varying degrees, which has been found to lead to a loss of blood quality resulting in both increased and decreased protein detection levels (37, 45, 46). The proteome has a wide degree of natural variation which must be considered in addition to pre-processing conditions when conducting studies that use high-throughput proteomics.

This pilot study was initiated to test protocols and parameters of a larger cohort study that is currently underway (17). This was a very small pilot conducted to ensure the research team was aware of potential limitations and risks to data outcomes, as no other published reports detailed how the SomaScan 7K assay would be impacted by the use of capillary blood (collected from Tasso+ devices) as well as the effects of pre-analytical variation on the proteome. Therefore, it was necessary to conduct this pilot prior to the initiation of the larger studies which use Tasso+ kits to collect blood for other, more diverse analyses. The small sample size and lack of replicates (due to cost restraints in this study) reduced robustness of the study. More participants and biological replicates would allow for a clearer definition of the impact of adverse pre-processing conditions on the proteome to identify what variation is truly due to simulated shipping conditions versus what is individual variation between participants. Temperatures above 37°C should also have been tested – initially a blood sample was collected from each participant using Tasso kits and stored at 55°C, but the blood was so visibly degraded it was not feasible or cost-effective to analyze it. More specific understanding of the impacts of time and temperature could be deciphered by collecting more samples at room temperature for different time periods at different time points (particularly, room temperature for 24 hours, or less-extreme high temperatures (i.e. 40°C) should be studied in the future. Despite this, it is clear that efforts to control pre-processing conditions are essential to minimize these impacts to get an accurate picture of the proteome using at-home blood collection kits and aptamer-based proteomics assays.

## Conclusions

This pilot study found that using capillary blood will result in a similar detected proteome to venous blood when analyzed using high-throughput proteomics if the samples are centrifuged and serum is frozen immediately. When processing delays are under 24 hours, samples can be stored at room temperature or shipped with an ice pack to keep the temperature low. When looking at very specific proteins or groups of proteins, rather than the overall proteome, utilizing mail-in kits may fall short, as a diverse range of specific proteins were impacted by adverse processing conditions, however, given the changes seen throughout the majority of the proteome, these tests may be most valuable when analyzing the proteome as a whole rather than individual proteins. In conclusion, the proteome of capillary and venous blood samples was found to be comparable using the SomaScan 7K assay and further studies are needed to determine impacts on individual protein expression levels.

## Data Availability

All data produced including SomaLogic raw data, processed data, and all code used for data analysis models, sorting of significant proteins into pathways, and data visualization are available online at GitHub: https://github.com/carolinescranton01/AtHomeBloodCollection-Proteomics_Analysis

## Contributors

**CS**: Conceptualization, methodology, experimentation, formal analysis, writing – original draft, and review & editing. **KKC**: Conceptualization, methodology, resources, funding acquisition, project administration, supervision, writing, and review & editing. **KPB**: Conceptualization, methodology, funding acquisition, writing, and review & editing. **EA**: Methodology, writing, and review and editing. **XS**: Methodology, formal analysis, and review & editing. **CMM**: Experimentation, writing, and review & editing. **DR**: Experimentation and review & editing. **VO**: Experimentation and review & editing.

## Abbreviations

PCA: principal components analysis
RFU: relative fluorescence units
L2FC: log 2-fold change
SST: serum separating tube
KEGG: Kyoto encyclopedia of genes and genomes

## Acknowledgements

The authors thank Jeff Buckthal, Alex Anaele, Kathryn Jenko, and Will Schwarzmann at Somologic for technical assistance in sample preparation, processing, and data analysis for the study. This study was supported by the National Institute of Diabetes and Digestive and Kidney Diseases (NIH’s NIDDK) award 1R01DK135483-01 provided to Drs. Kristen Pogreba-Brown and Kerry K. Cooper.

## Supplementary Materials

**Supplementary Table S1. Proteins with significantly altered detection for all samples**

**Supplementary Table S2. Proteins with significantly altered detection in every sample**

**Supplementary Table S3. Proteins with significantly altered detection in no samples**

**Supplementary Table S4. PERMANOVA and Kruskal-Wallis test results**

**Supplementary Table S5. Proteins with significantly altered detection in Model B**

**Supplementary Table S6. Proteins with significantly altered detection in Model C (Temperature)**

**Supplementary Table S7. Proteins with significantly altered detection in Model C (Time)**

**Supplementary Figure S1. SomaLogic Quality Statement**

Quality statement provided by SomaLogic, denoting the quality of the samples on arrival and other information regarding quality, calibration, and the assay.

**Supplementary Figure S2. Venn diagrams for individual variation**

Venn Diagrams for each participant (A through D, in panels A through D) which show the overlap in proteins which were detected at significantly different levels in each sample, compared to the capillary baseline samples.

**Supplementary Figure S3. Venn diagram of proteins altered in all samples per individual**

Venn diagram of the significantly impacted proteins in all of participant A, B, and D’s samples (participant C had no proteins altered in all samples compared to the baseline).

**Supplementary Figure S4. Significantly changed protein levels between capillary and venous samples of individual participants**

Four-panel bar graph of the proteins with significantly impacted detection for each of the participants when comparing the venous-collected sample to the capillary baseline. Four panels (A through D) correspond to participants A through D. Box-and-whisker plots are overlayed on the bars to show the spread of protein detection for each KEGG subcategory.

