## Supplementary Figure S1 for "Pilot Study to Determine the Efficacy, Feasibility, and Impact of Storage Conditions on At-Home Blood Collection Kits for Proteomic Studies"

somalogic™

### SomaScan® Assay Quality Statement

SomaLogic Operating Co., Inc.

2945 Wilderness Place

Boulder, CO 80301

**Study Name:** UAz-Cooper-Serum-Tasso Pilot Project-SOW9

**SomaLogic Study ID:** SS-2349076

**Sample Matrix:** Serum

**Date:** 2024-01-29

**Document Purpose:** Summary of results and key quality attributes from the analysis of samples with the SomaScan® proteomic platform performed by SomaLogic, Inc. This data is for Research Use Only (RUO).

#### Standardization

SomaScan assay data are first normalized using hybridization controls to mitigate variation within the run that comes from the readout steps: transfer to Agilent slides, hybridization, wash, and scan. This is followed by median signal normalization across pooled calibrator replicates within the run to mitigate within-run technical variation in the calibrator signal prior to use in scaling calculations. The set of ratios of the calibrator reference value to the median of calibrator replicates for each SOMAmer® reagent is calculated and decomposed into two terms: plate scale - the median ratio, and calibration scale - the recalculated set of scale factors, one for each SOMAmer reagent. Plate scale adjusts for overall signal intensity differences between runs. Calibration adjusts for SOMAmer reagent-specific assay differences between runs. Median signal normalization is performed using Adaptive Normalization by Maximum Likelihood (ANML) for specimen types and studies shown to be consistent with predefined population references or, alternatively, using median normalization to a study specific reference.

Acceptance criteria are shown below. Non-Core Matrices are often not subject to all data standardization procedures.

©2022 SomaLogic, Inc. SomaLogic, SomaScan, SOMAmer and associated logo are trademarks owned by SomaLogic Operating Co., Inc.

### Sample Normalization

##### Sample Summary

The total number of samples and controls are listed below, including the number that are passed or flagged by the quality criteria. Flagged samples are considered outlier samples and may differ due to biological or technical variables. Outliers can provide useful information about the sample quality, disease state, or study area; understanding the potential causes of outliers is crucial in deciding whether to exclude them from analysis.

| | Sample | QC | Buffer | Calibrator | Sum |
|---|---|---|---|---|---:|
| Total | 32 | 3 | 6 | 5 | 46 |
| PASS | 32 | 3 | 6 | 5 | 46 |
| FLAG | 0 | 0 | 0 | 0 | 0 |

##### Median Normalization Scale Factors

Median Normalization Scale Factors are median of the selected set of ratios for each sample of the SOMAmer reagent signal within dilution group to the population reference. Values are expected to be within the range of 0.4-2.5.

| Dilution Group | PASS | FLAG | TOTAL |
|---|---|---|---:|
| NormScale_20 | 32 | 0 | 32 |
| NormScale_0_005 | 32 | 0 | 32 |
| NormScale_0_5 | 32 | 0 | 32 |

##### ANML Fraction Used

ANML Fraction Used refers to the fraction of a dilution group for each sample that is between 2 population reference standard deviations of normal population reference value. Values between 2 population reference standard deviations are used to calculate a 'NormScale Factor' for a dilution group. If the ANML Fraction Used is less than 30%, the sample will be flagged; Greater than 30%, the sample will pass.

| Dilution Group | PASS | FLAG | TOTAL |
|---|---|---|---:|
| ANMLFractionUsed_20 | 32 | 0 | 32 |

©2022 SomaLogic, Inc. SomaLogic, SomaScan, SOMAmer and associated logo are trademarks owned by SomaLogic Operating Co., Inc.

| | | | |
|---|---|---|---|
| ANMLFractionUsed_0_005 | 32 | 0 | 32 |
| ANMLFractionUsed_0_5 | 32 | 0 | 32 |

### Calibration

##### Plate Scale

Plate Scaling is measured from the median of the set of calibrator reference ratios per plate, which results in a single scale factor to apply across the plate. It is typically associated with scanner intensity differences from plate to plate. Scale Factor Acceptance Criteria per plate is between 0.4-2.5.

| Plate | Acceptance Criteria | Plate Check | Value |
|---|---|---|---|
| PLT29473 | 0.4 - 2.5 | PASS | 0.96 |

### Quality Control Check

##### Calibrator Percent In Tails

Calibrator Percent In Tails refers to the percentage of plate calibration scale factors with values outside the expected range, 0.6 - 1.4.

| Plate | Alert Criteria | Plate Check | Value |
|---|---|---|---|
| PLT29473 | Less than 10% | PASS | 1.0 |

##### SOMAmers In Tails

SOMAmers In Tails refers to the cumulative number of SOMAmer reagents in the QC control with a ratio on any plate outside the accepted accuracy range, 0.8 - 1.2, when compared to the reference. Flagged SOMAmer reagents are typically retained for analyses since accuracy across all assay runs is a robust quality metric but is not a requirement for identification of meaningful biological signal.

| SOMAmer | Acceptance Criteria | ColCheck | Count |
|---|---|---|---|
| QC Ratio | 0.8 - 1.2 | PASS | 7420 |
| | | FLAG | 176 |
| | | TOTAL | 7596 |

©2022 SomaLogic, Inc. SomaLogic, SomaScan, SOMAmer and associated logo are trademarks owned by SomaLogic Operating Co., Inc.

##### QC Percent In Tails

QC Percent In Tails refers to the percentage of SOMAmer reagents in the QC Control that are outside the accepted accuracy range, 0.8 - 1.2, when compared to the reference.

| Plate | Acceptance Criteria | Plate Check | Percent |
|---|---|---|---|
| PLT29473 | Less than 15% | PASS | 2.3 |

##### Calibrator CVs

Percentiles for the distribution of Calibrator CVs on each plate is shown below.

| Cal Precision (%) | 10% | 50% | 90% |
|---|---|---|---|
| PLT29473 | 1.2 | 2.5 | 5.7 |

##### QC CVs

Percentiles for the distribution of QC CVs for all plates are shown below.

| QC Lot | nSamples | 10% | 50% | 90% |
|---|---|---|---|---|
| 200174 | 3 | 1.0 | 2.9 | 7.3 |

### Delivered Adats

Data are provided for multiple standardization stages to allow the end user to evaluate efficacy and bias of median normalization for individual samples. We recommend beginning analysis with the ADAT file that has been most normalized. For core matrices, which includes Human EDTA Plasma, Serum, or Urine, the most normalized file often ends with 'anmlSMP' which indicates ANML has been applied to Samples. Non-Core Matrices may be normalized with alternative methods, and ANML may not have been applied.

For ADAT File readers, please visit https://github.com/SomaLogic.

SS-2349076_v4.1_Serum.hybNorm.medNormInt.plateScale.calibrate.anmlQC.qcCheck.anmlSMP.adat

SS-2349076_v4.1_Serum.hybNorm.medNormInt.plateScale.calibrate.anmlQC.qcCheck.adat

©2022 SomaLogic, Inc. SomaLogic, SomaScan, SOMAmer and associated logo are trademarks owned by SomaLogic Operating Co., Inc.
