## Supplementary figures and images for "Pilot Study to Determine the Efficacy, Feasibility, and Impact of Storage Conditions on At-Home Blood Collection Kits for Proteomic Studies"

### Supplementary Figure S2

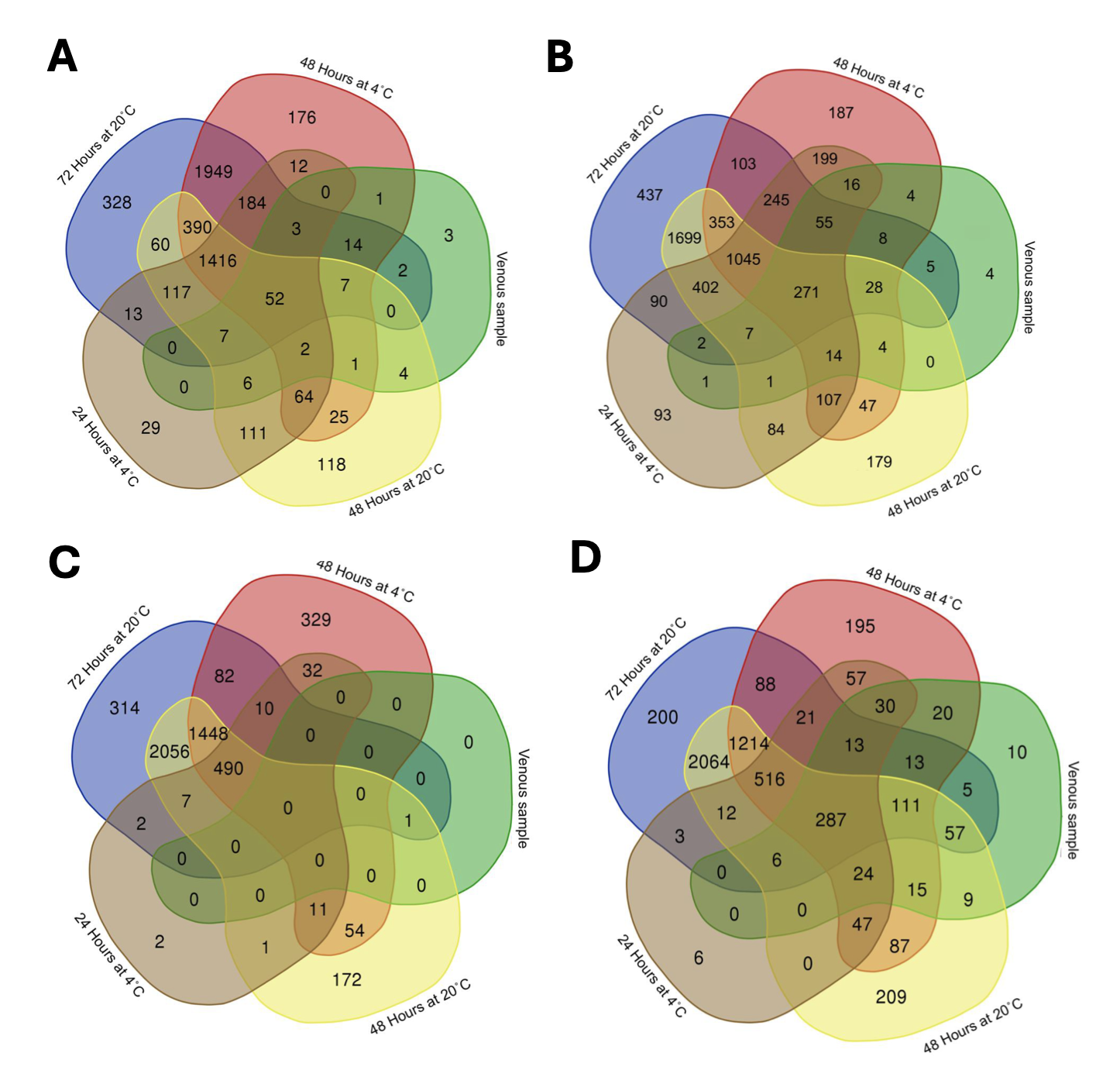

### Supplementary Figure S3

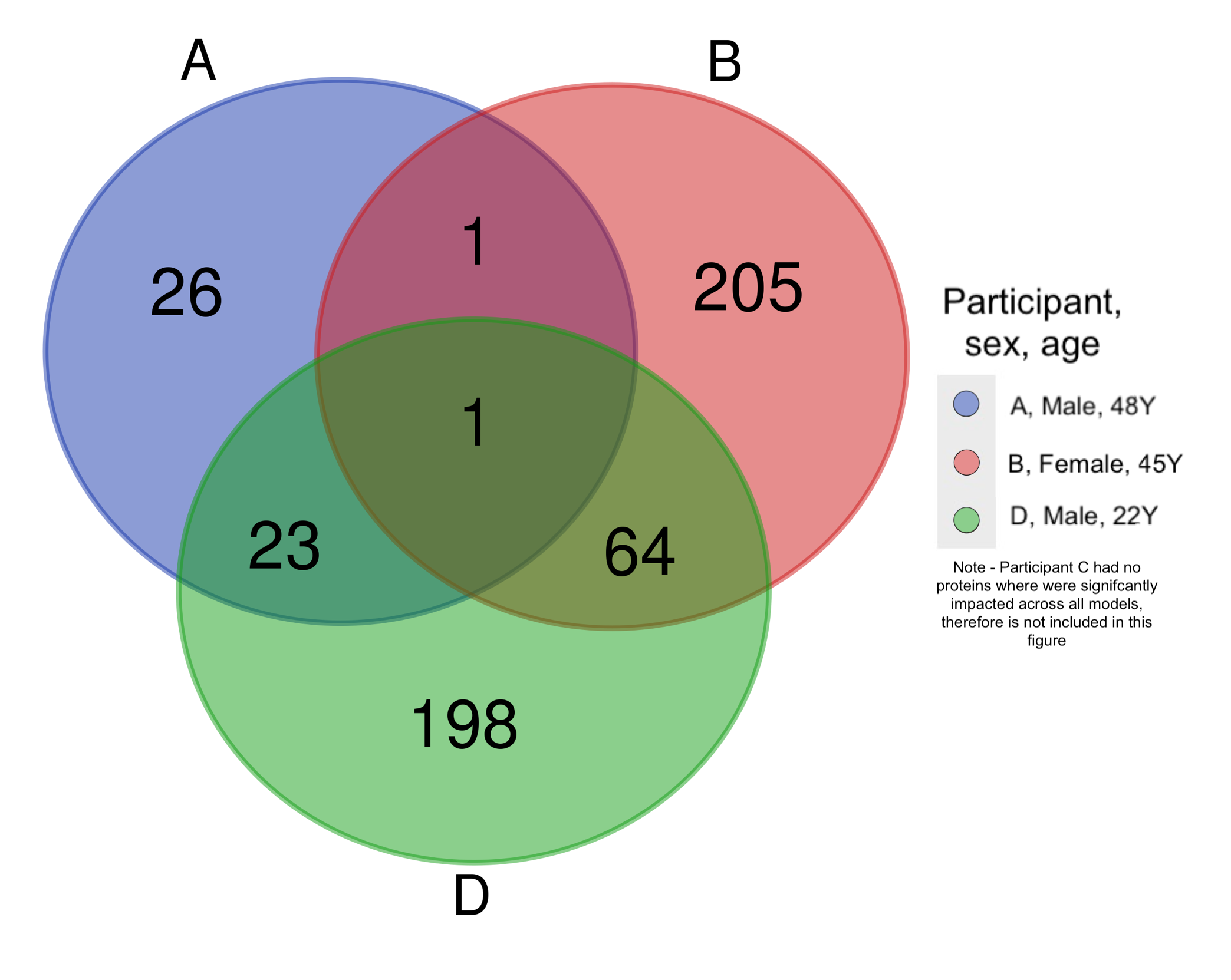

### Supplementary Figure S4

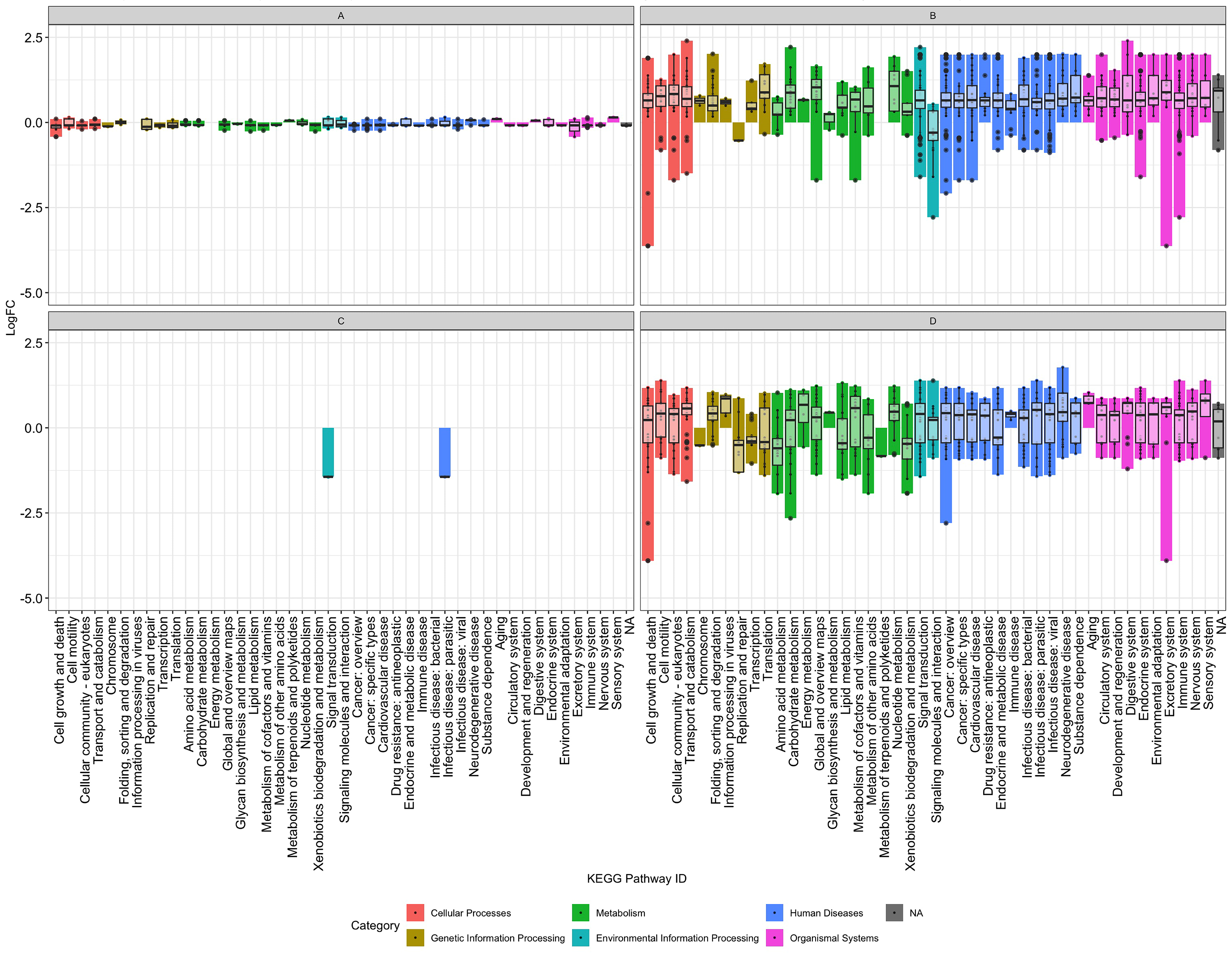
